# Effects of Exertion on PLR; A study of pupillary light reflex in rugby players before and after exercise

**DOI:** 10.1101/2024.10.10.24315265

**Authors:** W Davies, PM Middleton, I Buzytsky, Martha Ryan

## Abstract

**Aim:** To investigate the effects of physical exertion on pupillary light reflex (PLR) metrics in sports participants, with the aim of determining whether exercise-induced changes could potentially confound concussion assessment using PLR.

**Materials and methods:** Twenty-five adult rugby players underwent PLR assessment, using the MindMirror™ smartphone application, before and after a 30-55 minute rugby practice session or game. PLR metrics included latency, maximum and minimum pupil diameters, maximum constriction velocity, and pupil size correlation. Paired t-tests were used to compare pre- and post-exertion PLR metrics.

**Results:** No statistically significant differences were observed between pre- and post-exertion assessments in any of the measured PLR metrics (p > 0.05 for all metrics). The most notable, though still non-significant, change was in maximum constriction velocity (mean difference: 1.47 mm/s, p = 0.08 for pupil 1; 1.09 mm/s, p = 0.27 for pupil 2).

**Conclusion:** This study suggests that PLR metrics, as measured by a smartphone-based pupillometer, do not appear to be affected by rugby-related exertion. These findings support the potential use of PLR assessment in concussion screening protocols without the need to account for exercise-induced changes. However, further research with larger sample sizes and in full-contact situations is needed to confirm these results and their applicability in concussion assessment.

**Plain language summary:** Pupil size and reactivity have been critical components of the clinical assessment of patients with head injury and concussion for decades. Pupillary examination may provide critical information related to new or worsening intracranial pathology, and quantitative PLR examination is increasingly used outside of the hospital setting, particularly for concussion assessment in sports medicine. Exertion-dependent changes could confound valid measurement, however, and compromise the detection of concussion, so we investigated PLR assessment with the MindMirror^™^ smartphone application. We found no systematic differences in PLR variables between exertion and non-exertion groups.

**Tweetable abstract:** A novel smartphone-based AI tool did not reveal exertion-related differences in pupillary light reflex measurement in rugby players.

## Introduction

With an increased awareness of the risks associated with sports-related concussion, baseline preseason testing has become common amongst adolescent and young adult athletes [1]. Testing using tools such as SWAY or SCAT 6 are undertaken by athletes preseason, and then again after a suspected concussion. The preseason baseline testing is often performed in a resting state.

There is evidence to suggest that exertion has effects on the outcomes of these tests. These exertion-dependent changes could confound valid measurement, and therefore compromise the detection of concussion. Several studies have demonstrated exertion-dependent changes to neurocognitive performance, which seem to persist for up to 20 minutes after exertion ceases. [2,3,4] Given that some neurocognitive testing regimens may take around 15 minutes, this would suggest that reliably assessing a potentially injured athlete may take up to 35 minutes off the pitch.

The pupillary light reflex (PLR) refers to a stereotyped pupillary constriction, and subsequent dilation, in response to a sudden increase in perceived light; this reflex is thought to be mediated by autonomic signals. Previous studies in young athletes with mild traumatic brain injury have demonstrated stereotypical abnormalities of the PLR associated with concussion, [5,6] which makes the PLR a potential candidate for objective testing. Given that other tools for concussion testing appear to be significantly affected by exertion, we decided to investigate whether the PLR were similarly affected.

The MindMirror™ smartphone application is a novel, automated pupillometer using a smartphone and its camera and flash capacity to capture videos of the pupils after a light stimulus. These images, in the form of 30/60 FPS video, are uploaded to a cloud-based artificial intelligence processor. The pupils and irises are identified and measured, and the algorithm accounts for important confounders relevant to the video capture such as distance, blinking, and tremor by using a smoothing algorithm. The application captures a 7 to 10-second clip which is then analyzed to generate a PLR curve. By using a smartphone, the application is accessible to those wishing to undertake pitch-side testing of potentially injured athletes.

The purpose of this study was to investigate the effects of exertion on the PLR curve captured by the MindMirror™ application. We hypothesized that there would be significant differences in PLR parameters before and after exertion, which could have implications for the use of PLR in concussion assessment protocols.

## Materials and methods

### Participants and study design

The study received low-risk ethics exemption after discussion with the Noosa Dolphins rugby team governance board. The club provided the subjects for the study. All participants were given study information and signed consent forms for the capture and storage of pupillary images. Participants were eligible for the study if they were over 18 years of age and free from any significant ophthalmic disease. Medical histories and medication histories were obtained from the participants prior to them undertaking baseline testing.

A convenience sample of 25 subjects was recruited before a non-contact training session of the rugby team. Baseline PLR curves were obtained from all participants; subsequently, curves were re-collected from the individuals within 5 minutes of them ceasing vigorous exercise. The vigorous exercise consisted of a rugby practice session or a game of rugby, each lasting between 30 and 55 minutes. The initial part of the training session focused on skills, while the latter part was a non-contact game. This non-contact game format was chosen to create emotional stimulus and exertion levels similar to those experienced in a real game, but without any risk of concussion.

### Data Collection

Participant videos were captured in indirect lighting conditions using three smartphone models: iPhone 11, iPhone 15 Pro, and Pixel 8 Pro. The MindMirror™ application’s interface provides a framing screen to optimize image capture for processing. Three investigators were trained to use the MindMirror™ mobile app. Their training focused on holding the device steady, using correct and consistent lighting conditions, and capturing pre/post exercise videos in a continuous batch with no interruptions. Images were captured by one of the three investigators present at the pitch and uploaded to the cloud for processing.

The post-game/post-exertion videos were captured no later than 10 minutes after the game. All videos were captured at the same location and at the same time of day. They were then uploaded to MindMirror™ servers, where pupil/iris detections were run on them frame by frame.

Raw data output for each video contained several metrics pertaining to frame-by-frame diameters of participant’s pupils and irises, and whether the video was captured before or after exercise.

## Results

**Table 1:**
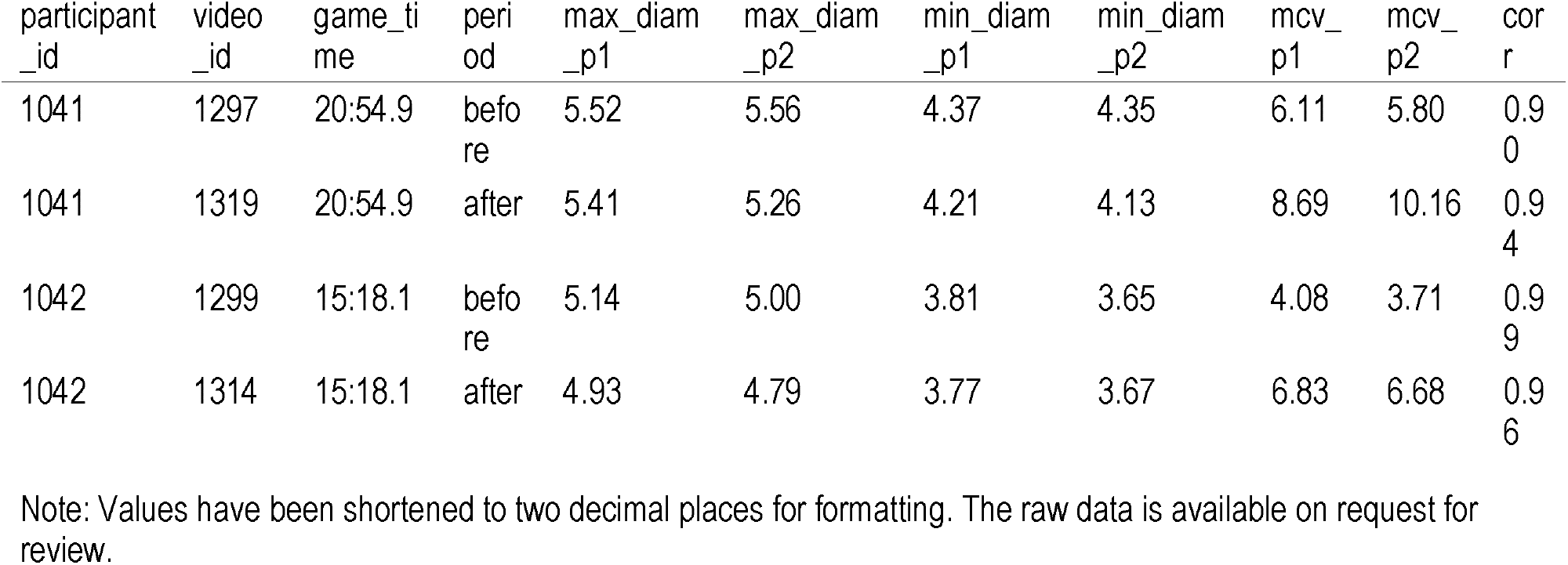
Raw PLR Data Sample.

**Table 2:** Raw PLR Metadata.

| Column Name | Description | Units |
| --- | --- | --- |
| participant_id | Unique identifier for each participant | - |
| test_id | Identifier for the specific test | - |
| video_id | Identifier for the video in which the frames are captured | - |
| frame_no | Frame number within the video | - |
| p1_score | Confidence score for pupil 1 detection | - |
| p1_diam | Diameter of pupil 1 | pixels |
| p2_score | Confidence score for pupil 2 detection | - |
| p2_diam | Diameter of pupil 2 | pixels |
| i1_score | Confidence score for iris 1 detection | - |
| i1_diam | Diameter of iris 1 | pixels |
| i2_score | Confidence score for iris 2 detection | - |
| i2_diam | Diameter of iris 2 | pixels |
| brightness | Brightness level of the frame | - |

### Data Analysis

Raw data was subsequently processed and analyzed. Data processing involved several steps:

1. Filtering of raw PLR data to include only entries with a valid score above a specified threshold.
2. Imputation of missing frame pupil diameter values using spline interpolation.
3. Merging of filtered data with additional metadata from participant information.
4. Smoothing of P1/2 diameters using the Savitzky--Golay filter with a window of 7 to conservatively eliminate detection noise.
5. Conversion of pupil diameters from pixels to millimeters, assuming an average human iris diameter of 11.6 mm.

Key metrics computed for both pupils (P1 and P2) included latency, maximum and minimum diameters, maximum constriction velocity (MCV), and recovery time.

**Table 3:**
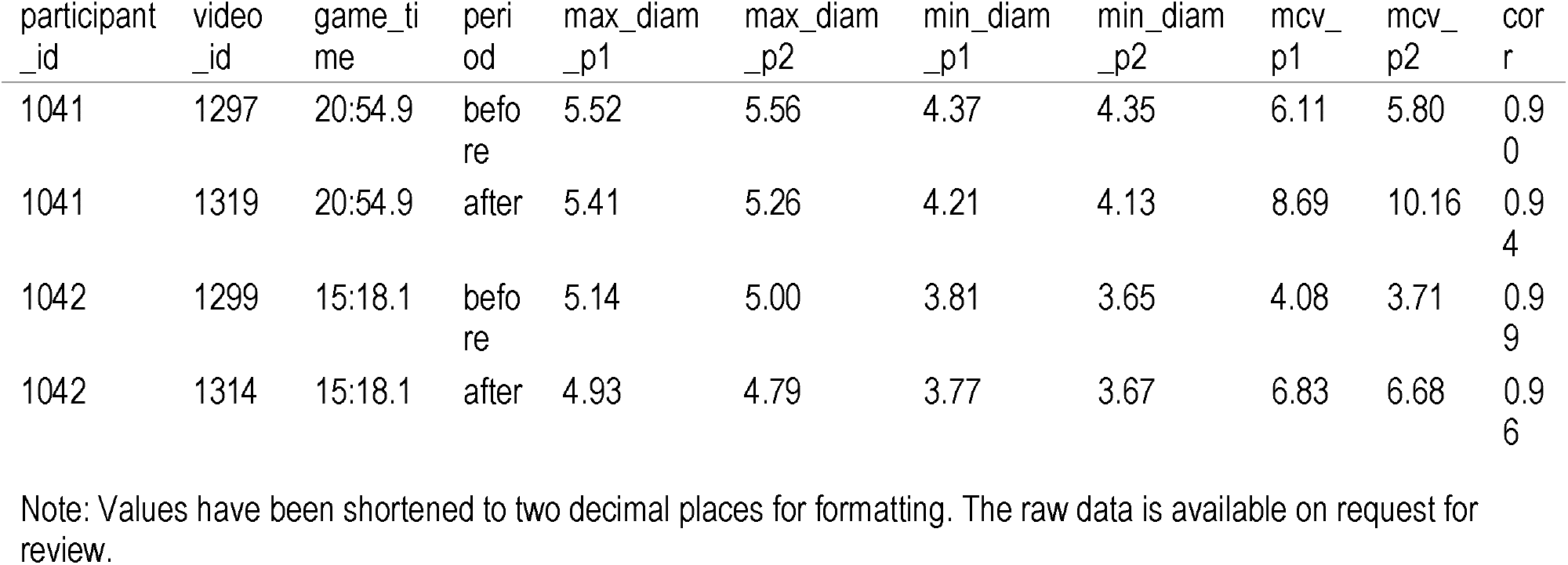
Key PLR Metrics.

**Table 4:** Key PLR Metrics Metadata Column Name Description.

| Column Name | Description | Units |
| --- | --- | --- |
| participant_id | Unique identifier for each participant | - |
| video_id | Identifier for the video in which the frames are captured | - |
| game_time | Timestamp of the game when the data was captured | - |
| period | Period of the game (before or after) | - |
| max_diam_p1 | Maximum diameter of pupil 1 | mm |
| max_diam_p2 | Maximum diameter of pupil 2 | mm |
| min_diam_p1 | Minimum diameter of pupil 1 | mm |
| min_diam_p2 | Minimum diameter of pupil 2 | mm |
| mcv_p1 | Maximum constriction velocity of pupil 1 | mm/s |
| mcv_p2 | Maximum constriction velocity of pupil 2 | mm/s |
| corr | Correlation between the measurements of pupil 1 and pupil 2 | - |

Statistical analysis was performed using paired t-tests for each PLR metric to compare the before and after periods. Mean differences and p-values were calculated for each metric, with statistical significance set at $p < 0.05$ .

**Table 5.** Non-Significant Differences.

| Metric | Mean Difference | p-value |
| --- | --- | --- |
| latency_p1 | 0.01 | 0.74 |
| latency_p2 | 0.00 | 0.95 |
| mcv_p1 | 1.47 | 0.08 |
| mcv_p2 | 1.09 | 0.27 |
| corr | -0.03 | 0.34 |
| recovery_time_p1 | 0.19 | 0.49 |
| recovery_time_p2 | 0.08 | 0.70 |
| variance_p1 | -0.07 | 0.24 |
| variance_p2 | -0.05 | 0.49 |

Note: There were no statistically significant differences observed when using a Savitzky--Golay filter with a window of 7 to eliminate some of the noise.

**Figure 1:**
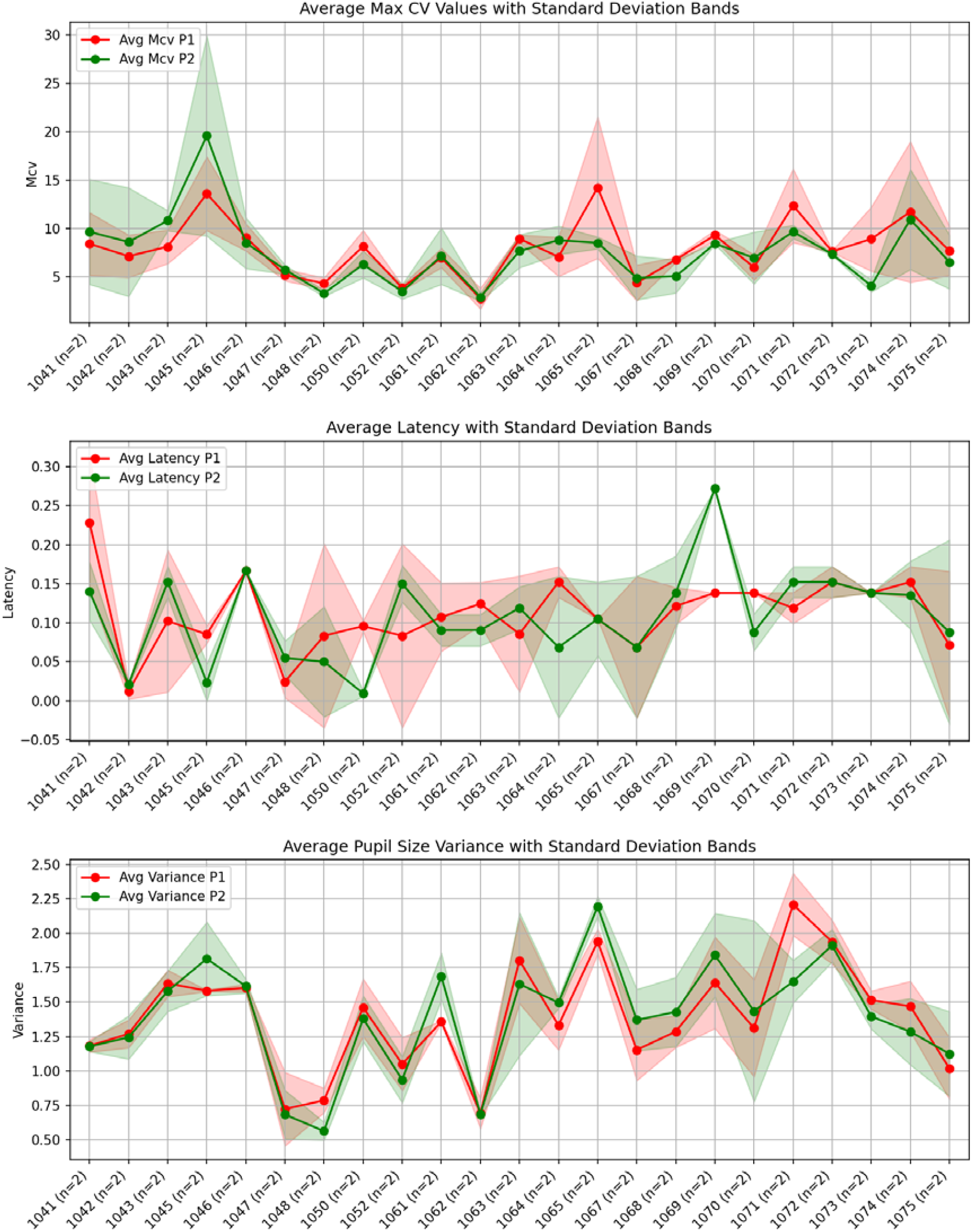
Key Pre / Post Exercise Metrics per participant, with STD bands.

## Discussion

Several important considerations and areas for future research emerge from this study.

### 1. Quality of Pupil Detection Models

While our study utilized YOLO (You Only Look Once) models for pupil detection, the field of machine learning is rapidly evolving. Future iterations of this research could explore additional techniques such as Mask R-CNN (Region-based Convolutional Neural Networks) or ensemble methods that combine multiple models. These approaches might yield even more accurate and robust pupil detection.

Moreover, the performance of these models could be significantly enhanced by training them on larger and more diverse cohorts. Specifically, including participants from various racial backgrounds and with different eye colors would improve the model’s generalisability. This is particularly important given the potential for variations in pupil detection accuracy across different iris pigmentations.

### 1. Age-Related Considerations

Our study focused on adult rugby players, but the effects of exertion on PLR might vary across different age groups. Future studies should consider including middle-aged and older adults, as well as adolescents. Age-related changes in autonomic nervous system function and pupillary response could influence how exertion affects PLR metrics. Understanding these age-related differences would be crucial for developing comprehensive, age-appropriate concussion assessment protocols.

### 1. Autonomic changes associated with level of athletic training

As an amateur rugby team, levels of fitness may have been subject to substantial variation between players. Moderate endurance training offers advantages such as enhanced parasympathetic activity and baroreflex sensitivity (BRS), along with a reduced level of sympathetic tone, however the effect of very intensive training on neural cardiovascular regulation is less known. Some studies have indicated that very intensive endurance training shifts cardiovascular autonomic modulation from a parasympathetic toward a sympathetic predominance [7]. The implication of altered blood pressure and vasomotor autonomic function has been little explored, leaving open the possibility of unpredictable impacts on measures such as the PLR.

### 1. Validation with FDA-Approved Devices

While our smartphone-based method shows promise, it is important to establish its accuracy and reliability against gold-standard measurements. Future studies should consider using FDA-approved pupillometer devices such as the NeurOptics NPi-200 or NPi-300 to establish a measurement ground truth. This comparison would not only validate our smartphone-based approach but also help quantify any discrepancies between the two methods.

### 1. Limitations and Future Directions

Several limitations of our study should be addressed in future research. First, our sample size was relatively small, and a larger cohort would provide more statistical power. Second, we focused on non-contact rugby activities to avoid potential concussions, but future studies should investigate PLR changes in full-contact situations, provided ethical considerations are adequately addressed.

Additionally, while we found no significant changes in PLR metrics post-exertion, subtle changes might become apparent with more sensitive measurement techniques or different analytical approaches. Future studies might consider more advanced statistical methods, such as machine learning algorithms, to detect nuanced patterns in PLR data.

Lastly, longitudinal studies tracking PLR changes over multiple training sessions and games could provide insights into cumulative effects of exertion and potential relationships with fatigue or overtraining.

## Conclusion

This study investigated the effects of rugby-related exertion on pupillary light reflex (PLR) metrics using the novel MindMirror smartphone application. Our analysis of 25 rugby players, using advanced smartphone models (iPhone 11, iPhone 15 Pro, and Pixel 8 Pro) and standardised data collection procedures, revealed no statistically significant differences in PLR metrics between pre- and post-exertion measurements.

Specifically, we found no meaningful changes in latency, maximum constriction velocity, correlation, recovery time, or variance of pupillary response after vigorous rugby practice or game play. These results suggest that the pupillary light reflex, as measured by our smartphone-based method, may be resilient to the effects of physical exertion in the context of rugby.

While our findings indicate that PLR metrics remain stable after exertion, it’s important to note that this study focused on non-contact rugby activities to avoid potential concussions. Further research is needed to determine whether these results hold true in full-contact situations or in cases of suspected concussion.

The stability of PLR metrics pre- and post-exertion suggests that this method could potentially be used for sideline concussion assessment without the need for extended rest periods. However, more extensive studies with larger sample sizes and diverse athletic populations are necessary to confirm these findings and establish the reliability of smartphone-based PLR assessment in sports medicine.

## Data Availability

All data produced in the present study are available upon reasonable request to the authors

## Statements and Declarations

- Non-financial interests: Dr Davies and Professor Middleton act as unpaid medical advisors to MindMirror inc.
- Financial interests: Mr. Buzytsky is a cofounder and CTO of MindMirror, Inc.

## Ethics approval

This study was performed in line with the principles of the Declaration of Helsinki, and under the SWSLHD Human Research Ethics Application (HREA) approval 2023/ETH00741: The Australian Pupillary Light Response Registry (APLRR). Informed consent was obtained from all volunteers.

